# Anticoagulants and Antiplatelets in COVID-19: Impact on Survival and Thromboembolism Development

**DOI:** 10.1101/2021.07.03.21254541

**Authors:** Thomas Salmon, Mitchell Titley, Zaid Noori, Mark Crosby, Rajiv Sankaranarayanan

**Affiliations:** Liverpool University Hospitals NHS Foundation Trust, Liverpool, United Kingdom; Liverpool Centre for Cardiovascular Science, University of Liverpool, United Kingdom

**Keywords:** COVID-19, thromboembolism, anticoagulation, antiplatelet, survival

## Abstract

**Background:** Higher rates of venous and arterial thromboembolism have been noted in coronavirus disease-2019 (COVID-19). There has been limited research on the impact of anticoagulant and antiplatelet choice in COVID-19.

**Methods:** This was a single-centre retrospective cohort study of 933 patients with COVID-19 infection presenting between 01/02/2020 and 31/05/2020. Survival time at 90 days post-diagnosis and thromboembolism development were the measured outcomes.

**Results:** Of 933 total patients, mean age was 68 years and 54.4% were male. 297 (31.8%) did not survive at 90 days. A Cox proportional hazards model analysis found no statistically significant relationship between anticoagulant or antiplatelet choice and survival (p<0.05).

57 (6.3%) developed thromboembolism. Antiplatelet choice was not shown to have a statistically significant relationship with thromboembolism development. Warfarin and direct oral anticoagulant (DOAC) use did not have a statistically significant impact on thromboembolism development (p<0.05). Therapeutic low-molecular-weight heparin (LMWH) use was associated with increased thromboembolism risk (Odds ratio = 14.327, 95% CI 1.904 – 107.811, p = 0.010).

**Conclusions:** Antiplatelet choice was shown to have no impact on survival or thromboembolism development in COVID-19. Anticoagulant choice did not impact survival or thromboembolism development, aside from LMWH. Therapeutic LMWH use was associated with increased risk of thromboembolism. However, it should be noted that the sample size for patients using therapeutic LMWH was small (n=4), and there may be confounding variables affecting both LMWH use and thromboembolism development. These findings should be repeated with a larger sample of patients using therapeutic LMWH with additional adjustment for cofounding variables.

## Introduction

Coronavirus disease 2019 (COVID-19) was first identified in December 2019, as a collection of atypical viral pneumonia cases centred around Wuhan, China. Over the following months COVID-19 transmission occurred on a global scale, leading to the World Health Organisation (WHO) to declare a COVID-19 pandemic on 11^th^ March 2020. Following this, the study of COVID-19 has become a priority worldwide. Knowledge of the clinical manifestations of the disease has developed significantly, leading to the understanding that infections can range from asymptomatic to severe and life threatening [1,2], and to the recognition that symptomatic patients can present with both respiratory and non-respiratory symptoms [3,4]. Crucially, it has been noted that COVID-19 infection carries an increased risk of thromboembolism [5–7], which has been associated with increased mortality [5].

Owing to the observed links between COVID-19 infection, thromboembolism, and mortality, there has been research into the impact of prophylactic or treatment dose anticoagulation on both mortality and on thromboembolism incidence in COVID-19 positive patients. Results of research on the impact of prophylactic or treatment anticoagulation on all-cause mortality have been mixed, with some finding no impact of anticoagulation (prophylactic or therapeutic) on mortality [8,9], whilst others note a statistically significant decrease in mortality in patients receiving prophylactic or therapeutic anticoagulation when compared with patients receiving no anticoagulation [10–12]. Furthermore, there are conflicting findings on whether higher doses of anticoagulation than typical prophylactic doses lead to decreases in mortality, with some finding no statistically significant difference between patients receiving prophylactic or therapeutic anticoagulation [11], whilst others have identified that the use of “intermediate doses”, defined as higher than prophylactic but lower than therapeutic, can lead to a statistically significant decrease in mortality when compared with prophylactic doses [13].

Conclusions of research on the impact of anticoagulant therapy on thromboembolism in COVID-19 positive patients has been similarly mixed. Some have found venous thromboembolic risk in critically-ill patients with COVID-19 infection to be significantly higher with prophylactic anticoagulation when compared with therapeutic anticoagulation [14], whereas some retrospective cohort studies have found no statistically significant difference in thrombosis development between groups receiving prophylactic anticoagulation, therapeutic anticoagulation, and intermediate doses [15,16]. However, much of this research is limited to patients admitted to critical care, or studied relatively small populations.

Whilst, as discussed, there has been significant study of the impact of therapeutic and prophylactic anticoagulation on both thromboembolic risk and mortality in COVID-19 positive patients, little research exists on the choice of anticoagulant. This study aims to investigate whether the choice of therapeutic anticoagulant has an impact on either thromboembolic risk or mortality in patients diagnosed with COVID-19 infection, assessing the differences between patients receiving warfarin, direct oral anticoagulants (DOACs), therapeutic low molecular weight heparin (LMWH), and those receiving prophylactic anticoagulation or no anticoagulation.

Additionally, patients hospitalised with COVID-19 have been shown to be at higher risk of arterial thromboembolism [17,18]. The number of studies on the impact of antiplatelet therapy in patients with COVID-19 infection is low in comparison with those on anticoagulation. Of the existing data, a recent meta-analysis found antiplatelet use had no significant impact on mortality in patients admitted with COVID-19 infection [19]. Despite this, it is theorised that antiplatelet agents may have a protective effect on patients with COVID-19 infection [20], and a recent prospective cohort study has identified a trend of decreased mortality in patients receiving aspirin, although this was not deemed statistically significant due to study size [21]. This study will aim to add to the current literature by assessing the impact of antiplatelet therapy on mortality and incidence of thromboembolism.

Finally, this study aims to add to the current literature on the impact of pre-existing cardiovascular disease on outcomes for COVID-19 patients. Patients with heart failure have been shown to be at higher risk of hospitalisation and death due to COVID-19 [22], and ischaemic heart disease has been associated with more severe COVID-19 infection [23]. Much has been noted of arrhythmia in COVID-19 patients as a clinical manifestation of the disease [24,25], but less research has been conducted on the impact of pre-existing arrhythmia. As such, this study aims to assess the impact of pre-existing arrhythmia on mortality and thromboembolism incidence, whist also assessing the impact of heart failure and ischaemic heart disease on the same outcomes.

## Methods

We performed a single centre retrospective cohort study analysing the impact of cardiovascular disease history, anticoagulant choice, and antiplatelet choice on survival and the incidence of thromboembolism in 933 patients presenting to a single hospital with PCR or imaging-proven COVID-19 infection between the dates of 01/02/2020 and 31/05/2020. We established a database of patients with PCR or chest/thoracic imaging proven COVID-19 infection based on imaging and microbiology performed through the Aintree University Hospital NHS Trust. We included patients who had imaging (CT thorax/chest or chest radiography) or PCR results reported between the dates 01/02/2020 to 31/05/2020. For included patients, we collected survival time at 90 days from the first COVID-19 positive PCR report or the first positive/probable COVID-19 imaging report. We also assessed whether patients were found to have thromboembolism on admission, or whether a thromboembolism was discovered at a later point during a patient’s admission.

We collected data on patient cardiovascular disease history and risk factors for thromboembolism development. We also reviewed each patient’s medication, collecting information on the use of and choice of antiplatelet and anticoagulant medication.

Statistical analysis was performed using SPSS version 27.0.1.0. Univariate chi-squared tests, t tests, and multivariate logistic regression testing was performed when analysing thromboembolism outcomes, and survival was analysed using a Cox proportional hazards model. All analysis used a p value of <0.05 for statistical significance.

### Inclusion criteria: Patients

We collected all patients who had a positive COVID-19 PCR test, or suggestive thoracic imaging (either CT chest/thorax, or chest radiography), reported between the dates of 01/02/2020 and 31/05/2020. This included patients who were inpatients within the hospital prior to 01/02/2020, in addition to patients presenting to the hospital during this period, regardless of admission status.

### Inclusion criteria: Diagnosis of COVID-19 on PCR swab

Between the dates 01/02/2020 and 31/05/2020, all nose and throat swabs tested for COVID-19 through PCR were reported electronically. We included all patients who had a positive swab result reported between 01/02/2020 and 31/05/2020. During these dates PCR testing for COVID-19 was only performed when there was a clinical suspicion of infection, and it was not hospital policy to swab all new patients admitted during this period.

### Inclusion criteria: Diagnosis of COVID-19 on imaging

During the period in question, all CT scans and chest radiography performed at Aintree University Hospital were reported by radiologists. All CT imaging reports and chest radiography reports were coded with a CVCX0, CVCX1, CVCX2, or CVCX3 as per the British Society of Thoracic Imaging (BSTI) guidelines [26]. The designation and description for each code can be found in table 1.

**Table 1.** BSTI Covid-19 imaging report codes

| Code | Description |
| --- | --- |
| VCX0 | Normal.<br>COVID-19 not excluded. Correlated with RT-PCR. |
| VCX1 | Classic/Probable COVID-19.<br>Lower lobe and peripheral predominant multiple opacities that are bilateral (>> unilateral). |
| VCX2 | Indeterminate for COVID-19.<br>Does not fit Classic or Non-COVID-19 descriptors. |
| VCX3 | Non-COVID-19.<br>Pneumothorax / Lobar pneumonia / Pleural effusion(s) / Pulmonary oedema / Other |

All patients with thoracic CT and chest radiograph imaging reported between the dates 01/02/2020 and 31/05/2020 with CVCX1 coding were included in our database.

### Exclusion criteria

Duplicate entries or patients with missing electronic records were excluded from the study population. Of 979 cases returned through the inclusion criteria, 46 cases were excluded due to missing/incomplete electronic records or being duplicate entries.

### Outcome: Survival at 90 days

Once the list of included patients was established, we collected survival time for all included patients. Survival time was measured in days from the time of the first COVID-19 positive PCR swab or imaging report, and was capped at 90 days. Any patient with survival above 90 days from the time of their first positive PCR swab or imaging report was recorded as having a survival time of “above 90 days”.

The upper limit of 90 days survival was chosen as data collection started in September 2020, just over 90 days after 31/05/2020 (the date of the last included PCR/imaging reports).

### Outcome: Thromboembolism

For all included patients we collected information on whether new thromboembolism was identified on initial presentation, and whether thromboembolism was identified at a later point during a patient’s admission. Only patients with thromboembolism definitively proven on imaging were included.

2 patients had known thromboembolism on presentation. One had a longstanding mural thrombus, and one had a chronic abdominal aortic thrombus. For data analysis, these patients were recorded as not having thromboembolism on imaging, as we aimed to assess the association between COVID-19 and new thromboembolism, rather than pre-existing thromboembolic disease.

### Data collection: cardiac history, risk factors

Cardiovascular history was collected for all included patients. This included history of ischaemic heart disease, heart failure, or arrhythmia. We also collected information on the thromboembolic risk factors of smoking status, oral hormonal contraceptive or hormonal replacement therapy use, immobility, recent surgery, active or previous cancer, history of COPD, and whether each patient had previously developed a thromboembolism.

This information was collected from electronic patient notes, electronic service records, and paper notes recorded at the time of the patient’s most recent presentation to hospital.

### Data collection: Anticoagulation, antiplatelets

Finally, we reviewed each included patient’s medication and recorded whether or not a patient was prescribed antiplatelet or anticoagulant medication. For patients prescribed antiplatelets the choice of antiplatelet and whether the patient was taking single or dual antiplatelet therapy was recorded. For anticoagulated patients, the choice of DOAC, therapeutic dose LMWH, or warfarin was recorded. Medication lists were obtained from electronic prescribing records, or when not available (such as for patients who were not admitted to hospital) medication lists recorded during clerking or summary care records were used.

### Statistical analysis

All statistical analysis was performed using SPSS version 27.0.1.0. For survival at 90 days, analysis of all variables was carried out using a cox proportional hazards model. A p value of <0.05 was considered statistically significant.

For thromboembolism development, univariate analysis of the impact of categorical risk factors was assessed using the chi-squared test. For the one quantitative risk factor, age, t test was used. The risk factors found to be statistically significant in univariate analysis were then included in multivariate logistic regression analysis. As above, for all analysis a p value of <0.05 was considered statistically significant.

## Results – Survival at 90 Days

### Survival at 90 days: General information

Of 933 total patients included, 297 (31.8%) did not survive to 90 days after their first positive COVID-19 PCR swab or COVID-19 suggestive imaging. The mean age of those surviving at 90 days was 63 years (SD 16.9), whereas the mean age of the non-survival group was 78 years (SD 12.1 years). 516 (55.3%) of the whole population was male, rising to 62.3% (n=185) in the non-survival group. 52% (n=331) of the surviving group were male. The incidence of ischaemic heart disease was higher in the non-survival group versus the survival group (26.6%, n=79, versus 18.2%, n=116), as was the incidence of heart failure (16.2%, n=48, versus 12.9%, n=82), and previous thromboembolic disease (7.4%, n=22, versus 6.6%, n=42). The incidence of atrial fibrillation/flutter was almost doubled in the non-survival group versus the survival group (23.9%, n=71 versus 12.9%, n=82), with almost half (46.4%) of patients with atrial fibrillation/flutter not surviving at 90 days. However, as shown by the cox proportional hazards analysis below, this was not deemed to be independent of other variables.

Current smoking habit was found to be higher in the non-survival group (30.0%, n=89, versus 21.1%, n=134). It is important to note that smoking status was absent in the records of 333 (35.7%) of the 933 total patients. Decreased mobility was found to be higher in the non-survival group versus the survival group (97%, n=288, versus 84.1%, n=535). The non-survival group had higher incidence of active cancer (15.2%, n=45, versus 5.8%, n=37) and previous cancer history (10.4%, n=31, versus 3.8%, n=24).

The incidence of chronic obstructive pulmonary disease was found to be lower in the non-survival group versus the survival group (6.8%, n=63, versus 9.8%, n=91), as was the history of surgery within the past 12 weeks or during admission (1.5%, n=14, versus 4.9%, n=46).

See Table 2 for full population characteristics.

**Table 2.**
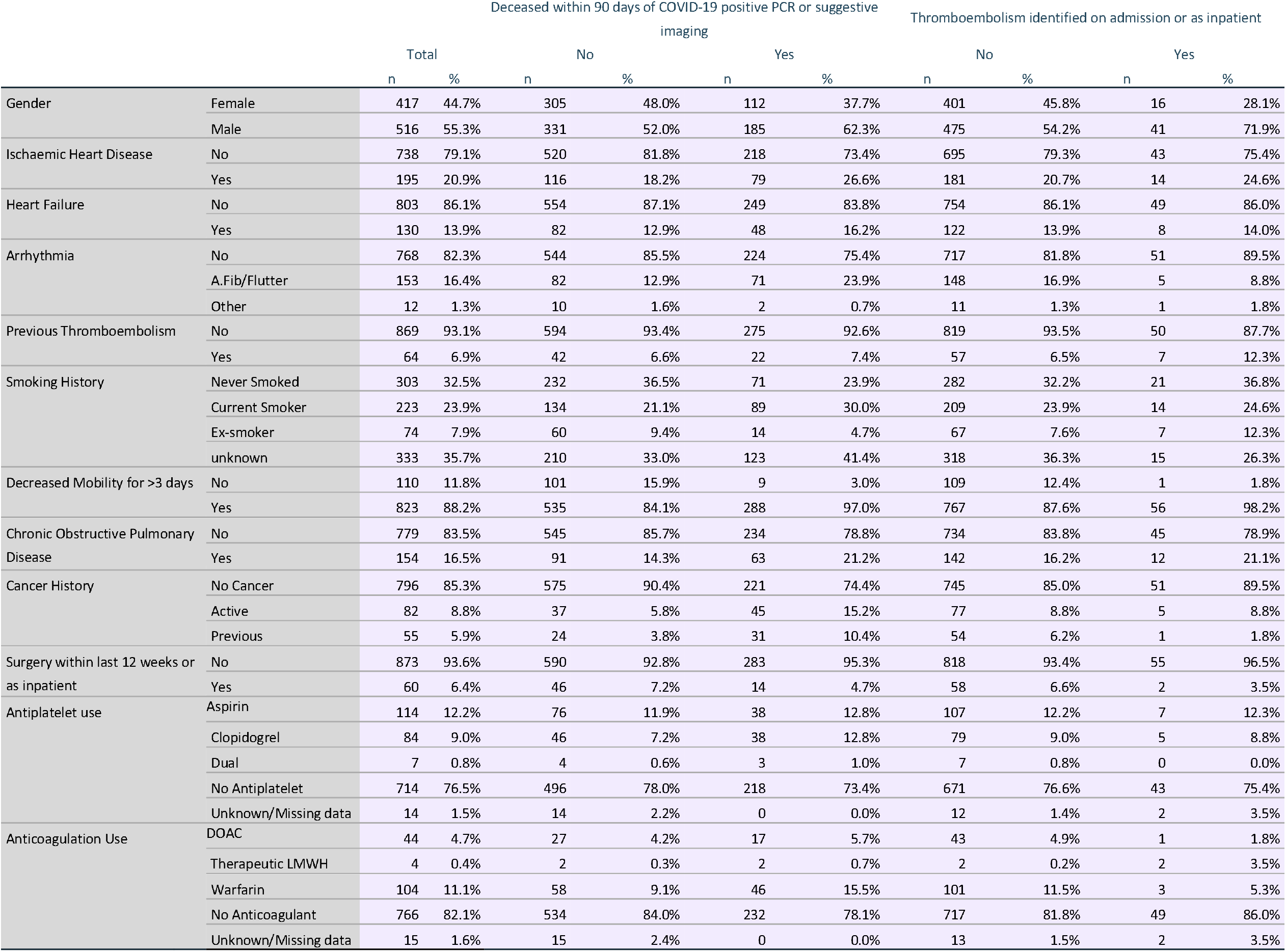
Population Characteristics

### Survival at 90 days: Regression analysis using Cox proportional hazards model

A Cox proportional hazards model analysis was performed using all risk factors, with survival time at 90 days as the outcome variable. As shown in Table 3, age, male sex, current smoking habit, decreased mobility, recent surgery, and active or previous cancer were found to be statistically significant independent factors affecting survival in patient with COVID-19 infection (p<0.05). All aside from recent surgery were found to increase risk of mortality, with decreased mobility (hazard ratio 2.264, 95% CI 1.150 - 4.457) having the largest effect, followed by previous cancer (hazard ratio 1.802, 95% CI 1.216 - 2.672), active cancer (hazard ratio 1.656, 95% CI 1.186 - 2.311), current smoking habit (hazard ratio 1.463, 95% CI 1.050 - 2.038), male sex (hazard ratio 1.432, 95% CI 1.121 - 1.828), and finally age (hazard ratio 1.050, 95% CI 1.040 - 1.061).

**Table 3:** Cox proportional hazards model for survival at 90 days in patients with COVID-19 infection

|  | Hazard Ratio | 95.0% Confidence Interval for Hazard Ratio |  | p value |
| --- | --- | --- | --- | --- |
|  |  | Lower | Upper |  |
| Age | 1.050 | 1.040 | 1.061 | **<0.001** |
| Sex |  |  |  |  |
| Female | Reference |  |  |  |
| Male | 1.432 | 1.121 | 1.828 | **0.004** |
| Ischaemic Heart Disease | 0.954 | 0.776 | 1.429 | 0.738 |
| Heart Failure | 0.954 | 0.678 | 1.341 | 0.785 |
| Arrhythmia History |  |  |  |  |
| No Arrhythmia | Reference |  |  | 0.271 |
| Atrial Fibrillation/Flutter | 1.181 | 0.727 | 1.355 | 0.501 |
| Other Arrhythmia | 0.358 | 0.088 | 1.463 | 0.153 |
| Previous Thromboembolism | 0.852 | 0.537 | 1.350 | 0.495 |
| Smoking History |  |  |  |  |
| Never Smoked | Reference |  |  | **0.012** |
| Current Smoker | 1.463 | 1.050 | 2.038 | **0.025** |
| Ex-Smoker | 0.684 | 0.373 | 1.253 | 0.219 |
| Smoking History unknown | 1.336 | 0.991 | 1.801 | 0.057 |
| Decreased Mobility | 2.264 | 1.150 | 4.457 | **0.018** |
| Chronic Obstructive Pulmonary Disease | 1.246 | 0.916 | 1.693 | 0.161 |
| Cancer History |  |  |  |  |
| No Cancer History | Reference |  |  | **<0.001** |
| Active Cancer | 1.656 | 1.186 | 2.311 | **0.003** |
| Previous Cancer | 1.802 | 1.216 | 2.672 | **0.003** |
| Recent Surgery (Within 12 weeks or During Admission) | 0.422 | 0.245 | 0.728 | **0.002** |
| Antiplatelet medication use |  |  |  |  |
| No antiplatelet medication | Reference |  |  | 0.736 |
| Aspirin alone | 0.786 | 0.539 | 1.145 | 0.210 |
| Clopidogrel alone | 1.036 | 0.714 | 1.501 | 0.853 |
| Dual antiplatelets | 0.949 | 0.289 | 3.114 | 0.931 |
| Anticoagulation Medication |  |  |  |  |
| No therapeutic anticoagulation | Reference |  |  | 0.804 |
| Warfarin | 0.767 | 0.445 | 1.323 | 0.340 |
| DOAC | 0.840 | 0.449 | 1.570 | 0.585 |
| Therapeutic LMWH | 1.511 | 0.332 | 6.875 | 0.593 |

Recent surgery, defined as within 12 weeks prior to admission, or as an inpatient during the COVID-19 related admission, was shown to be associated with an improved survival (hazard ratio 0.422, 95% CI 0.245 - 0.728).

Ischaemic heart disease, heart failure, previous thromboembolism, and chronic obstructive pulmonary disease were all found to have no statistically significant impact on survival independent of other variables. Pre-existing arrhythmia was also found to have no statistically significant independent impact on survival, despite only 54.4% of patients with atrial fibrillation or flutter surviving at 90 days.

Regarding antiplatelet use, survival was highest in those using no antiplatelets, followed by aspirin alone, and then clopidogrel (Figure 1). Survival was lowest in those taking dual antiplatelets. However, as noted in Table 3, there was no statistically significant relationship between antiplatelet choice and survival at 90 days.

**Figure 1:**
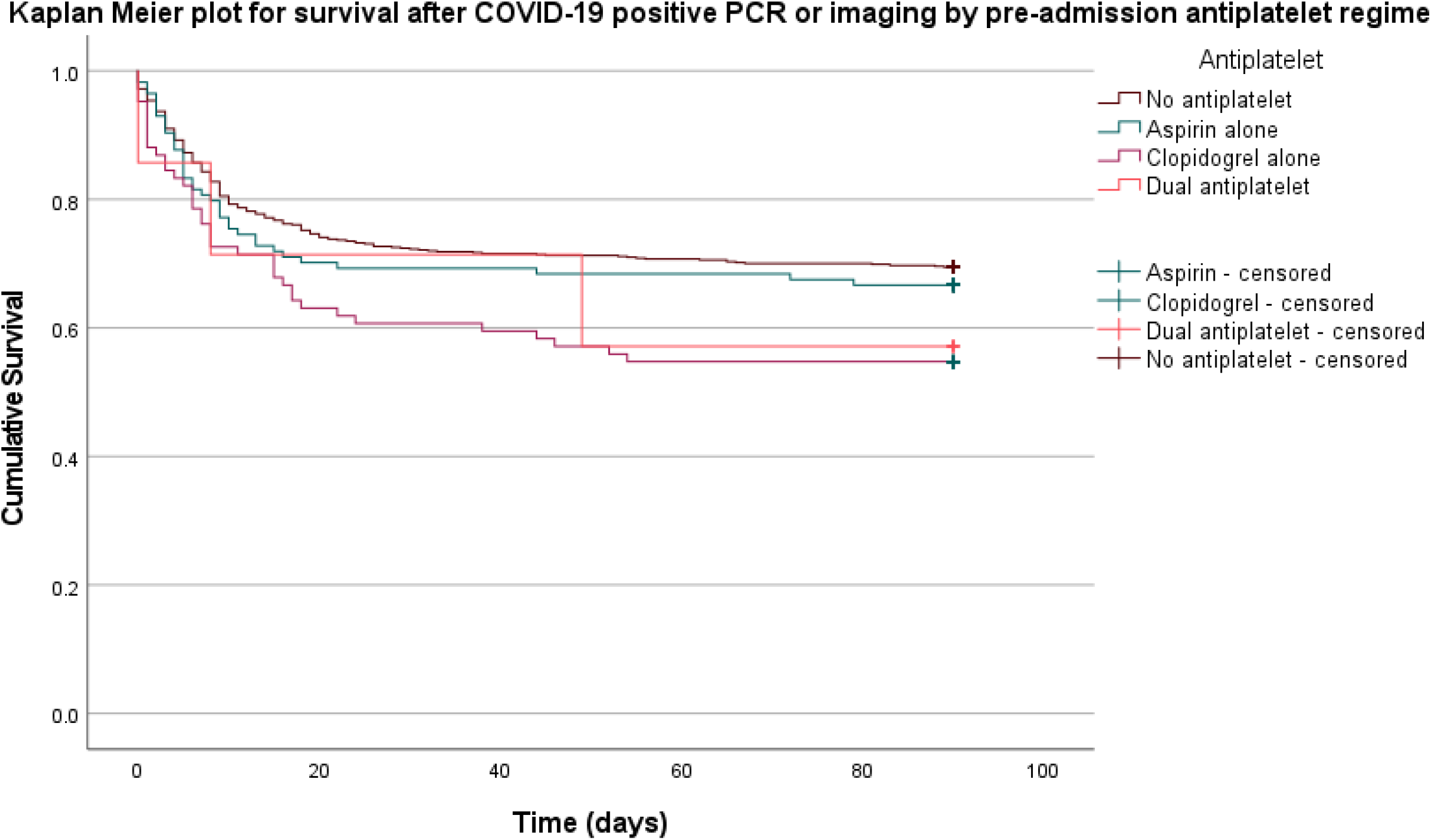
Survival at 90 days in COVID-19 positive patients by pre-admission antiplatelet regime

Between groups on different treatment doses of anticoagulation medication, survival was highest in those taking no treatment dose anticoagulation, followed by those taking DOACs, followed by those taking warfarin (Figure 2). Those on treatment dose LMWH had the lowest survival, although the sample size was small (n=4). As with antiplatelet choice, there was no statistically significant independent relationship between the use of warfarin, DOAC, LMWH or no therapeutic anticoagulation and survival at 90 days (Table 3).

**Figure 2:**
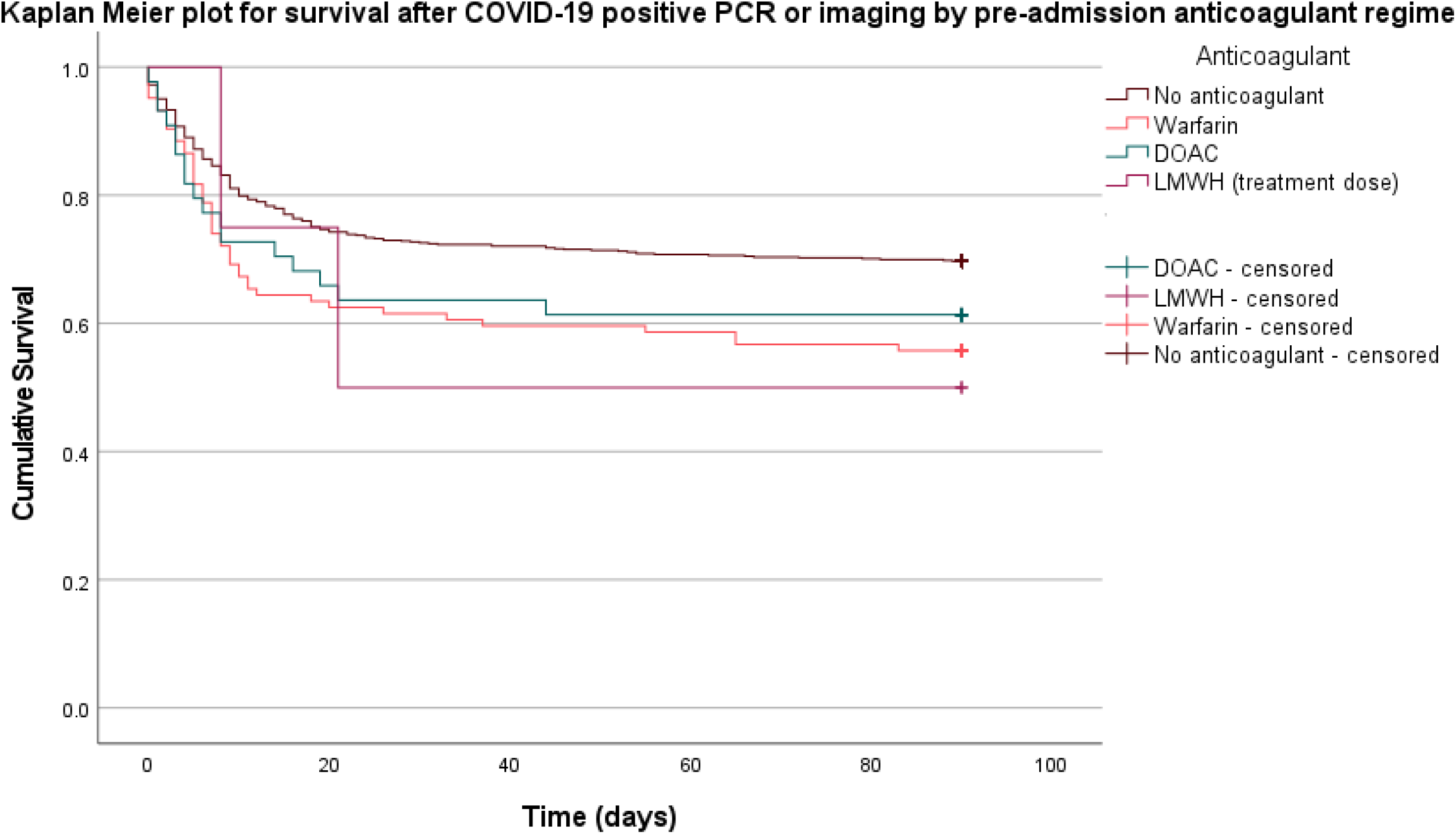
Survival at 90 days in COVID-19 positive patients by pre-admission anticoagulation regime

## Results – Thromboembolism

### General information: Thromboembolic disease

Of 933 total patients, 57 (6.3%) were found to have thromboembolism on imaging either on first presentation or as an inpatient. The mean age of those with thromboembolism was 65 years (SD 13.3), compared with a mean age of 68 years (SD 17.1) in those without thromboembolism (Table 5). As stated above, the study population was 55.3% male, rising to 71.9% in the thromboembolism group (n=41) versus 54.2% in the non-thromboembolism group (n=475). The incidence of ischaemic heart disease was higher in the thromboembolism group versus the non-thromboembolism group (24.6%, n=14, versus 20.7%, n=181), as was the incidence of previous thromboembolism (12.3%, n=7, versus 6.5%, n=57), decreased mobility (98.2%, n=56, versus 87.6%, n=767), chronic obstructive pulmonary disease (21.1%, n=12, versus 16.2%, n=142)

The incidence of heart failure was relatively equal between the thromboembolism and non-thromboembolism group (14.0%, n=8, versus 13.9%, n=122), as was the case with current cancer (8.8%, n=5, versus 8.8%, n=77). The incidence of atrial fibrillation was lower in the thromboembolism group (8.8%, n=5, versus 16.9%, n=148), as was the incidence of recent surgery (3.5%, n=2, versus 6.6%, n=58) and previous cancer (1.8%, n=1, versus 6.2%, n=54).

The incidence of all categories of smoking history (never smoked, ex-smoker, current smoker) was higher in the thromboembolism group, offset by lower incidence of those with unknown smoking history. As such, this result is of no significance.

There was little difference in the use of antiplatelets between the two groups, with no significant difference in the prevalence of aspirin, clopidogrel, dual antiplatelet or no antiplatelet use between the thromboembolism group and the non-thromboembolism group. 7 patients in the study were taking dual antiplatelets, none of which developed thromboembolism. However, this was not found to be statistically significant in univariate analysis, as shown below.

The prevalence of warfarin (5.3%, n=3, versus 11.5%, n=101) and DOAC (1.8%, n=1, versus 4.9%, n=43) use was lower in the thromboembolism group. The prevalence of no anticoagulant use was higher in the thromboembolism group (86.0%, n=49, versus 81.8%, n=717), as was therapeutic LMWH use (3.5%, n=2, versus 0.2%, n=2).

### Thromboembolism: Univariate analysis

Univariate analysis of categorical risk factors using the chi-squared test found male sex, decreased mobility, and anticoagulation use to be statistically significant factors when considering the development of thromboembolism in patients with COVID-19 infection (p<0.05) (Table 4). Of note, the comorbidities of ischaemic heart disease, heart failure, arrhythmia, chronic obstructive pulmonary disease, and active or previous cancer were found to have no statistically significant relationship with thromboembolism development in the setting of COVID-19 infection. Additionally, the risk factors of smoking, previous thromboembolism, and recent surgery were found to have no statistically significant relationship with thromboembolism development. Finally, the use of either single or dual antiplatelet therapy was found to have no statistically significant impact.

**Table 4:**
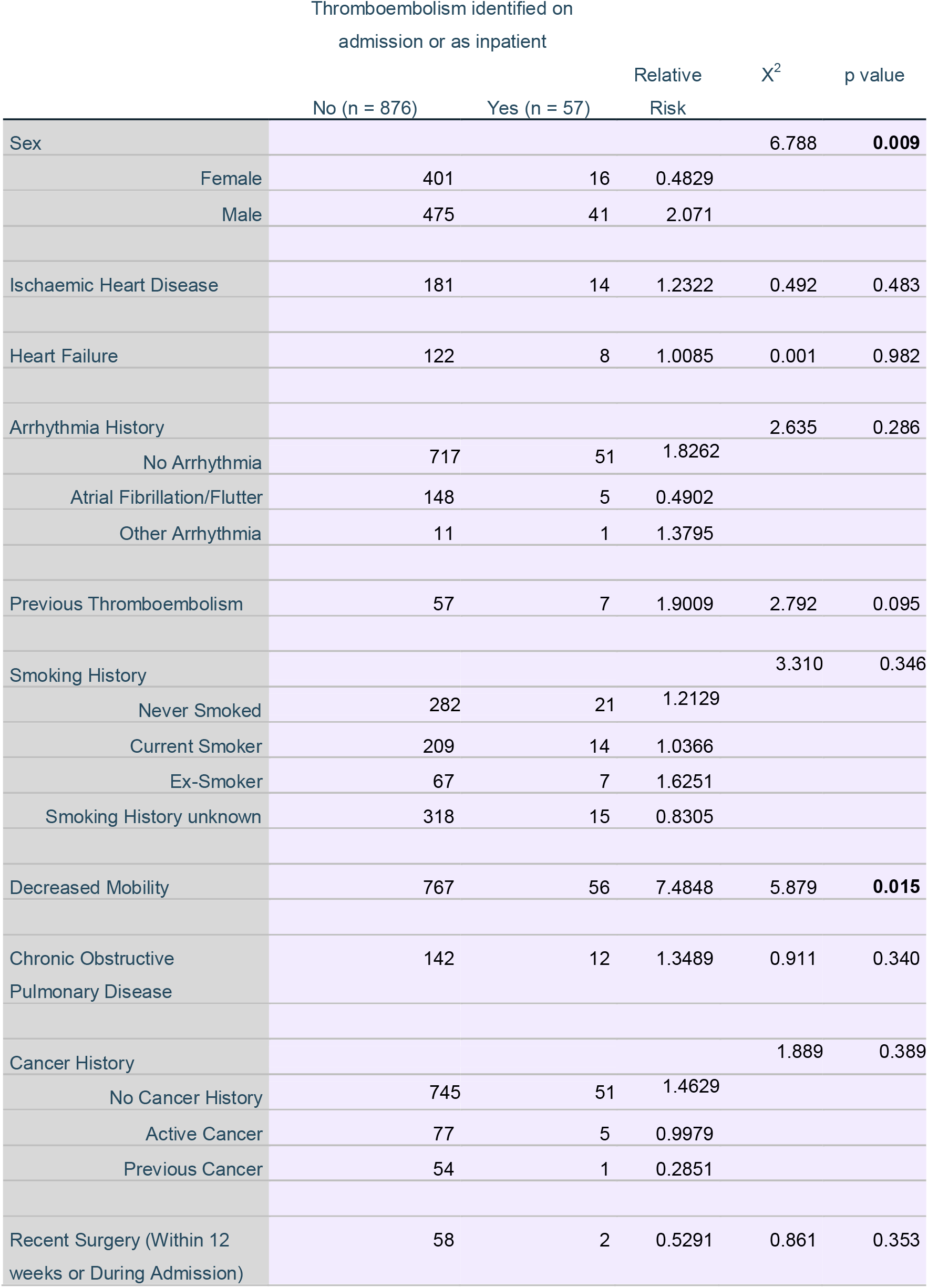
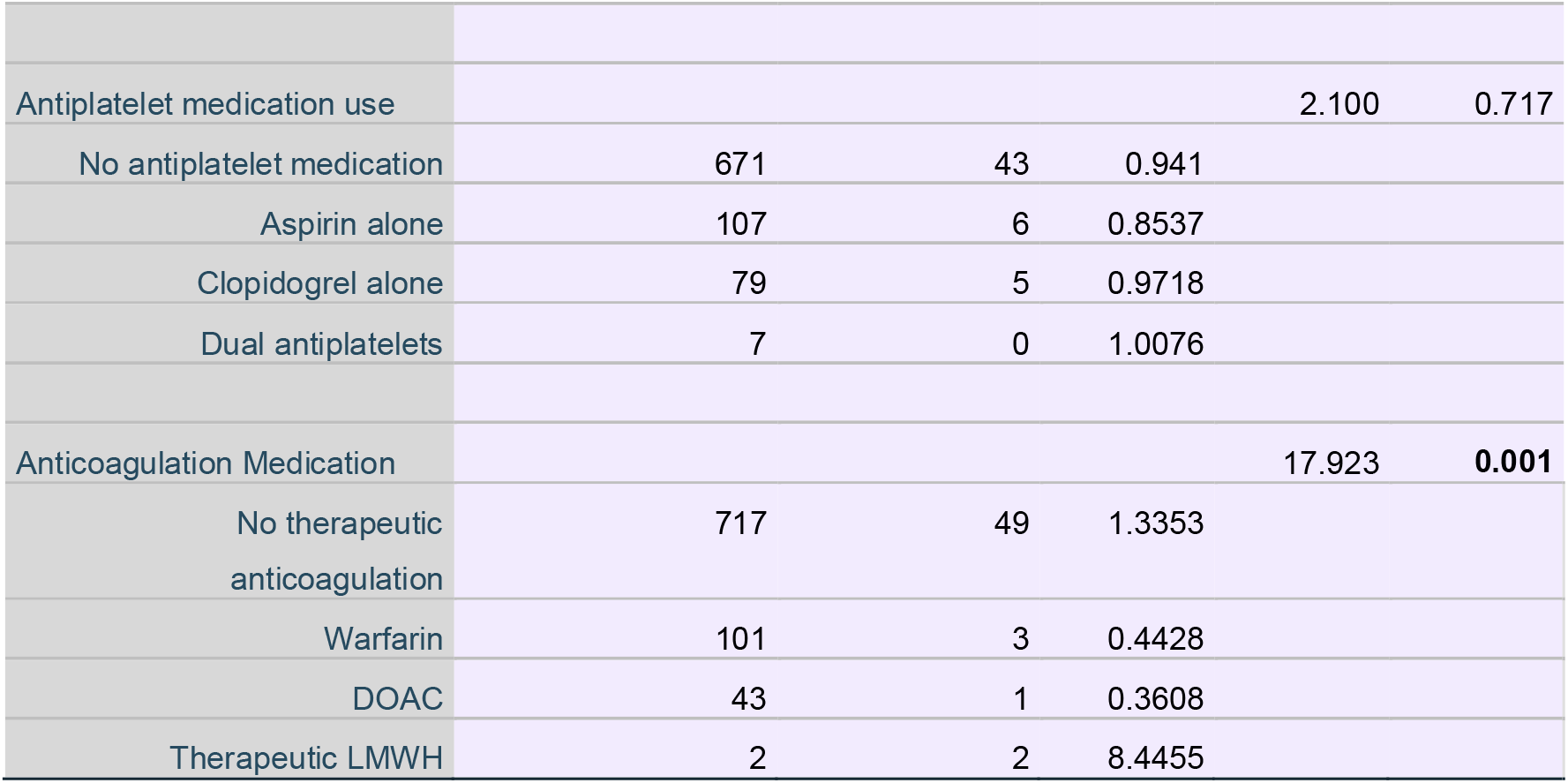
Univariate analysis of categorical risk factors for thromboembolism development in patient with COVID-19 infection using chi-squared test

**Table 5:** Age characteristics of COVID-19 patients with and without thromboembolism

| Thromboembolism on<br>imaging |  | n | Mean age | Std. Deviation | Std. Error Mean |
| --- | --- | --- | --- | --- | --- |
| No |  | 876 | 68.15 | 17.158 | .580 |
| Yes |  | 57 | 65.30 | 13.283 | 1.759 |

As shown in Table 6, univariate analysis of age using the 2-tailed independent samples t-test found no statistically significant difference in age between the thromboembolism group and the non-thromboembolism group (mean 65.30 years, SD 13.283, versus mean 68.15 years, SD 17.158, p=0.218).

**Table 6:** Univariate analysis of age as a risk factor for thromboembolism development in patients with COVID-19

|  | t | Sig. (2-tailed) | Mean<br>Difference | Std. Error<br>Difference | 95% Confidence Interval<br>of the Difference |  |
| --- | --- | --- | --- | --- | --- | --- |
|  |  |  |  |  | Lower | Upper |
| Equal variances<br>assumed | 1.232 | 0.218 | 2.855 | 2.317 | -1.692 | 7.402 |

### Thromboembolism: Multivariate regression analysis

Multivariate logarithmic regression analysis was performed using the statistically significant variables, namely sex, decreased mobility, and anticoagulant use. As shown in Table 7, male sex, decreased mobility, and therapeutic LMWH use were found to have statistically significant relationships with the development of thromboembolism in patients with COVID-19 infection (p<0.05), all 3 correlating with an increased risk of thromboembolism development. The strongest impact is attributed to therapeutic LMWH use (Odds ratio = 14.327, 95% CI 1.904 – 107.811), although it must be noted that the sample size for patients using therapeutic LMWH was very small (n=4). The next largest impact was due to decreased mobility (Odds ratio = 8.408, 95% CI 1.137 - 62.172), followed by male sex (Odds ratio = 2.063, 95% CI 1.128 – 3.774). DOAC use and warfarin use were not found to have a statistically significant independent association with thromboembolism development in patients with COVID-19 infection.

**Table 7:**
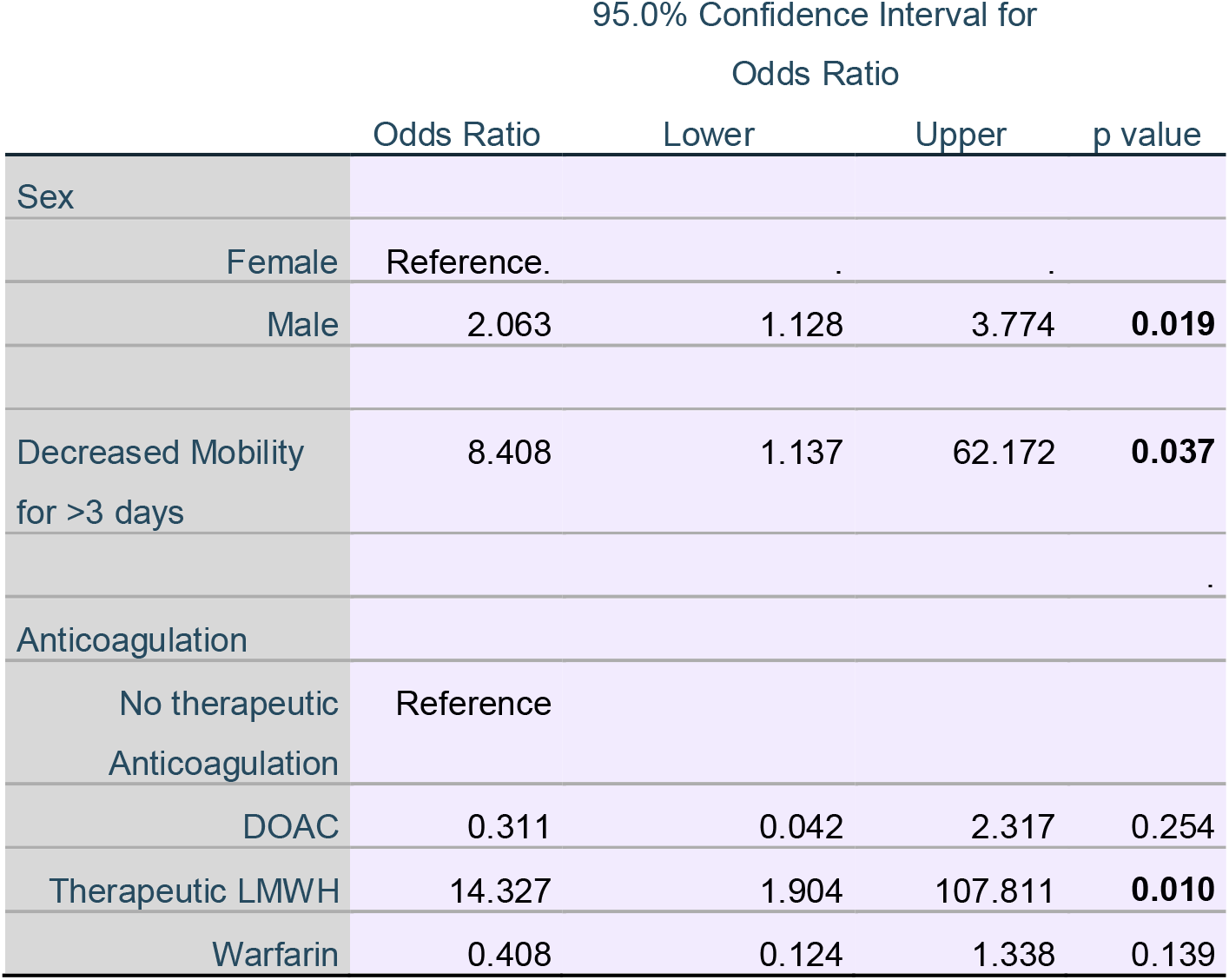
Multivariate logistic regression analysis of risk factors for thromboembolism development in patients with COVID-19 infection

## Discussion

### Discussion - Survival

The findings of this study on the impact of anticoagulant use are in line with other findings that therapeutic anticoagulation had no statistically significant impact on survival in patients with COVID-19 infection [8,9]. This study adds to current knowledge by demonstrating that the choice of anticoagulant does not change this finding – warfarin, DOAC, and therapeutic LMWH use were all found to have no significant impact on survival.

Survival was shown to be highest in patients not taking antiplatelets at the time of presentation and highest in those taking dual antiplatelets, showing that the theorised protective mechanisms [20] of antiplatelet use in COVID-19 positive patients did not have clinical ramifications in this population. The fact that the choice of antiplatelet, including no antiplatelet and dual antiplatelet use, was found to have no statistically significant impact on survival independent of population characteristics and cardiovascular comorbidities is in line with recent meta-analysis [19], and suggests a confounding factor may be responsible for the higher survival in patients not using antiplatelets. An obvious consideration would be pre-existing cardiovascular disease or cardiovascular risk, a common cause for antiplatelet use. Further investigation into a potential role for antiplatelet medication in COVID-19 is already underway, with aspirin being the subject of numerous ongoing trials [27]. Of note, the large RECOVERY trial (Randomised Evaluation of COVID-19 Therapy) is currently evaluating the impact on mortality of a single daily dose of 150mg until discharge [28], and the REMAP-CAP (Randomised Embedded Multi-factorial Adaptive Platform Trial for Community-Acquired Pneumonia) is aiming to assess the impact of aspirin, clopidogrel, ticagrelor and prasugrel [29].

Regarding population characteristics, it is unsurprising that age, male sex, active cancer, and current smoking habit were associated with poorer survival, as this has been demonstrated by numerous other studies [9,30–35]. The observation that survival was increased in those undergoing surgery during admission or in the 12 weeks prior to presentation is of note, but is potentially due to a better baseline of health. Whilst this study accounted for cardiovascular history and comorbidities associated with thromboembolic disease, it must be noted that an exhaustive list of patient comorbidities and lifestyle factors was not collected. A more complete analysis of a patient’s health as part of pre-operative assessment could act as a selection tool, meaning that the group undergoing surgery likely had a minimum level of fitness due to a selection bias that was not applied to the group not undergoing surgery.

From a cardiovascular history perspective this study found that pre-existing arrhythmia, heart failure, and ischaemic heart disease had no impact on survival at 90 days in patients with COVID-19 infection. Despite only 54.4% of the cohort with a history of atrial fibrillation surviving at 90 days, the relationship between pre-existing atrial fibrillation and survival was not shown to be independent of population characteristics and comorbidities. This suggests the existence of pre-existing atrial fibrillation in this population was an indicator of poorer overall health, rather than an independent factor contributing to COVID-19 related mortality.

### Discussion – Thromboembolism

The observation that treatment dose LMWH use at the time of presentation was associated with increased thromboembolism risk needs to be interpreted in context. The population size of patients using therapeutic LMWH on admission was very small, consisting of only 4 patients. Of these, 3 had active cancer, 3 had atrial fibrillation, and 2 had previously developed venous thromboembolism. By the mean Geneva risk score for venous thromboembolism in these 4 patients was 7.25 of a possible 30, suggesting they had a high baseline risk of developing venous thromboembolism [36]. Whilst the incidence of venous thromboembolism in patients using LMWH was 50%, the small sample size and potential for multiple confounding variables indicates that this result should be interpreted with caution.

The groups using warfarin, DOACs, and no therapeutic anticoagulation were of a good size, and as such more weight can be lent to the outcomes observed in these populations. The observation that there is no statistically significant difference in thromboembolism development between patients using warfarin, DOACs, and those not taking therapeutic anticoagulation is in line with some other cohort studies [15,16]. It does not align with the findings of numerous studies conducted in critically-ill patients [14,37–39], which found decreased incidence of thromboembolism in patients using therapeutic anticoagulation. This is likely a reflection of the differing severity of illness between study populations. As such, therapeutic anticoagulation may have a role in preventing COVID-19 associated thromboembolism in patients admitted to critical care, but this study found no benefit of warfarin or DOAC use in preventing thromboembolism in all patients presenting with COVID-19 infection.

Antiplatelet use was found to have no statistically significant relationship with thromboembolism development. This is in keeping with recent meta-analysis [19]. The role of clopidogrel is the subject of COVID-PACT (Prevention of Arteriovenous Thrombotic Events in Critically-Ill COVID-19 Patients), a randomised controlled trial of 750 patients investigating the impact of clopidogrel use on thromboembolism development with and without treatment dose anticoagulation [40]. This, alongside other ongoing trials [27], may find a role for antiplatelet therapy in the management of critically-ill COVID-19 patients. Despite multivariate analysis finding no statistical significance, it is of note that no patients taking dual antiplatelet therapy at the time of presentation developed thromboembolism. The sample size was small (n=7), and a statistically significant impact may be found with a larger sample size. At the time of writing, there are no ongoing clinical trials of dual antiplatelet therapy for thromboembolism prevention in COVID-19 [27]. There is, however, a current clinical trial of dipyridamole and aspirin co-administration in COVID-19, although the measured outcome is recovery and mortality rather than thromboembolism development [41].

The correlation between male sex and thromboembolism development in COVID-19 patients is in line with a recent meta-analysis [42]. Decreased mobility has been long-recognised as a risk factor for venous thromboembolism, thus the association with thromboembolism in this study is unsurprising.

### Limitations of study

As noted previously, the population sizes for groups taking dual antiplatelets (n=7) and therapeutic LMWH (n=4) were small. As such, findings related to these treatment groups should be interpreted with caution until studied with larger populations. Regarding dual antiplatelets, it is notable that no patient on dual antiplatelet therapy developed thromboembolism, although this was not deemed to be statistically significant in multivariate analysis. This warrants further study with a larger population. As mentioned previously, there are no current clinical trials of dual antiplatelet therapy for thromboembolism prevention in COVID-19 [27].

It is also important to note that the dose of antiplatelet medication taken was not recorded. The vast majority were taking 75mg aspirin daily, 75mg clopidogrel daily, or 75mg of each. Whilst the impact of higher dose antiplatelet regimes in COVID-19 certainly warrant investigation, a retrospective cohort study is not a suitable study design due to the underlying indications for higher dose aspirin or clopidogrel. Patients on higher doses will likely have suffered recent ischaemic stroke or acute coronary syndrome, and as such the impact of antiplatelet therapy on mortality and thromboembolism risk will be overshadowed by the risk attributable to these recent events. There are no ongoing large scale randomised control trials of the impact of higher dose antiplatelet medication in COVID-19 [27]. On a smaller scale, the CAM-Covid-19 trial is assessing the impact of 325mg aspirin given four times daily, alongside 0.6mg colchicine 12-hourly and 10mg montelukast daily, with a sample size of 34 patients [43].

Regarding LMWH, the observation that patients using LMWH were at higher risk of developing thromboembolism needs to be cautiously interpreted due to small sample size (n=4), and poor clinical health. As discussed previously, 75% of this group had active cancer and 50% had previously developed thromboembolism. Repeated study with a larger sample size and a matched cohort is suggested to assess whether this finding is truly independent of cohort characteristics and comorbidities.

## Conclusion

In all patients presenting to a single centre with COVID-19 infection, antiplatelet choice was shown to have no impact on survival at 90 days or thromboembolism development. With regards to anticoagulation, there was no statistically significant difference in survival at 90 days and thromboembolism development for groups using warfarin, DOACs, or no therapeutic anticoagulation. LMWH was associated with increased risk of thromboembolism, but sample size was very small (n=4), 75% of which had existing malignancy and 50% of which had developed thromboembolism in the past. Pre-existing ischaemic heart disease, arrhythmia, and heart failure were all found to not be associated with increased risk of thromboembolism or decreased survival at 90 days.

## Data Availability

Available on request from author.

## Abbreviations

COVID-19: Coronavirus disease 2019
WHO: World Health Organisation
DOAC: Direct Oral Anticoagulant
LMWH: Low Molecular Weight Heparin
PCR: Polymerase Chain Reaction
BSTI: British Society of Thoracic Imaging
A.Fib: Atrial Fibrillation
CI: Confidence Interval
SD: Standard Deviation

